# Inverse association between dietary fiber intake and asymptomatic intracranial atherosclerotic stenosis in older adults: The YAHABA Study

**DOI:** 10.64898/2026.03.29.26349674

**Authors:** Takashi Yamaguch, Ryo Itabashi, Hiroshi Akasaka, Eisuke Hirai, Masahiro Kudo, Naoki Ishizuka, Tetsuya Maeda

## Abstract

**Background:** Intracranial atherosclerosis is a major cause of ischemic stroke. Asymptomatic intracranial atherosclerotic stenosis (ICAS) represents a subclinical and potentially modifiable stage preceding ischemic stroke, yet the nutritional factors associated with asymptomatic ICAS remain poorly defined. This study aimed to identify dietary factors associated with asymptomatic ICAS in community-dwelling older adults.

**Methods:** This cross-sectional, population-based study included 962 Japanese adults aged ≥65 years from the Yahaba Active Aging and Healthy Brain study, conducted in Yahaba town, Japan, between July 2016 and July 2017. Asymptomatic ICAS was defined as ≥50% intracranial arterial stenosis evaluated by magnetic resonance angiography (MRA) without a history of stroke or transient ischemic attack. All participants underwent dietary assessment using a food frequency questionnaire. We examined the association between nutritional factors and ICAS using multivariable logistic regression models with adjustment for age, sex, hypertension, dyslipidemia, diabetes mellitus, body mass index, smoking, and alcohol use.

**Results:** After excluding 43 participants who could not undergo brain MRI, 2 with inadequate vascular imaging on MRA, 57 with a history of stroke, and 10 with missing data, 850 participants were included in the analysis. The mean age was 73.4 ± 6.5years, and 52% were female. ICAS was identified in 135 participants (15.9%) on MRA. Participants in the highest quartile of dietary fiber intake had significantly lower odds of ICAS than those in the lowest quartile (OR, 0.45; 95% CI, 0.26–0.80) after adjusting for vascular risk factors. Participants in the highest quartile of potassium intake also had lower odds of ICAS than those in the lowest quartile (OR, 0.49; 95% CI, 0.27–0.89). When dietary fiber and potassium were simultaneously included in the multivariable analysis as continuous variables, neither remained significant, with moderate collinearity (variance inflation factor, 4.16).

**Conclusions:** Higher dietary fiber intake was inversely associated with asymptomatic ICAS among community-dwelling older Japanese adults. Potassium intake also showed an inverse association, although this relationship was less consistent after accounting for collinearity with dietary fiber.

## 1. Introduction

Intracranial atherosclerosis is a major cause of ischemic stroke,^1, 2^ particularly in Asian populations, in whom its prevalence and disease burden are greater than in Western countries.^3^ In addition to causing symptomatic stroke, intracranial atherosclerotic disease may exist for prolonged periods without overt neurological manifestations.^4, 5^

Asymptomatic intracranial atherosclerotic stenosis (ICAS), defined as luminal narrowing of intracranial arteries not associated with prior ischemic stroke or transient ischemic attack (TIA), represents this subclinical stage of the disease. Accumulating longitudinal evidence suggests that asymptomatic ICAS is associated with an increased risk of future vascular and cerebrovascular events^4–6^ and may also be linked to higher long-term mortality.^5, 7^

Traditional vascular risk factors, including hypertension,^8, 9^ dyslipidemia,^10, 11^ and diabetes mellitus,^9, 12^ are well-established determinants of asymptomatic ICAS. However, potentially modifiable lifestyle factors beyond conventional risk markers have received less attention. In particular, the role of nutritional factors in the development of intracranial atherosclerosis remains incompletely understood. Dietary exposures are increasingly recognized as important determinants of cerebrovascular health^13^. Evidence from systematic reviews supports protective associations for higher dietary fiber intake^14^ whereas higher sodium intake and a higher sodium-to-potassium ratio have been associated with increased stroke risk.^15^ Nevertheless, most prior studies have focused primarily on clinical stroke outcomes or extracranial atherosclerosis, and data specifically addressing nutritional determinants of asymptomatic ICAS are scarce.

Given the high prevalence of ICAS in older Asian populations and its relevance as an early manifestation of ischemic stroke due to intracranial atherosclerosis, clarification of the association between dietary factors and asymptomatic ICAS could have implications for the prevention of cerebrovascular disease in community-dwelling Japanese older adults. Accordingly, the present study aimed to identify nutritional factors associated with asymptomatic ICAS among community-dwelling older Japanese adults using magnetic resonance angiography (MRA). By focusing on an asymptomatic population, we sought to elucidate modifiable dietary behaviors that may help prevent the development and progression of intracranial atherosclerosis before the onset of ischemic stroke or TIA.

## 2. Materials and methods

### 2.1. Subjects

We included participants from the Yahaba Active Aging and Healthy Brain (YAHABA) study, established in 2016 in Yahaba, a town in a rural area of Iwate Prefecture, Japan. The YAHABA study, a community-based prospective cohort study aimed at clarifying the risk factors and etiology of dementia, cerebrovascular disease, and movement disorders, is being conducted in collaboration with the Japan Prospective Studies Collaboration for Aging and Dementia.^16^ Details are described elsewhere.^16, 17^ This cross-sectional analysis was conducted using baseline data, including MRA and blood samples, collected between July 2016 and July 2017. This study was conducted in accordance with the Declaration of Helsinki. The protocol was approved by the Ethics Committee of Iwate Medical University (HGH28-12, HG2020-017), and written informed consent was obtained from all participants.

## Data collection

Detailed information on medical history, current medications, drinking, and smoking status was obtained using self-administered questionnaires, and medication status was determined by consulting participants’ medication records. Hypertension was defined as a blood pressure of ≥140/90 mmHg and/or current treatment with antihypertensive agents. Resting blood pressure was measured three times using an automated device in the sitting position after 5 minutes of rest, and the average of the three measurements was used in the analyses. Standard laboratory measurements, including serum lipid profiles (low-density and high-density lipoprotein cholesterol), glycated hemoglobin, and serum creatinine, were obtained at baseline. Estimated glomerular filtration rate (eGFR) was calculated using the equation proposed by the Japanese Society of Nephrology. Diabetes mellitus was defined as a random blood glucose concentration of ≥200 mg/dL, hemoglobin A1c (HbA1c) of ≥6.5%, and/or current treatment with antidiabetic medication. Dyslipidemia was defined based on the 2022 guidelines of the Japan Atherosclerosis Society as any of the following: low-density lipoprotein cholesterol ≥ 140 mg/dL, high-density lipoprotein (HDL) cholesterol < 40 mg/dL, triglycerides ≥ 150 mg/dL, or non-HDL cholesterol ≥ 170 mg/dL.

### 2.2. Brain imaging and definition of ICAS

The multi-slab 3D time-of-flight MRA was performed under one of the following imaging parameters: 1) Achieva 1.5T (Philips Healthcare, Netherlands), repetition time/echo time (TR/TE) 23/6.91 ms, field of view (FOV) 200 × 200 mm, slice thickness 1.6 mm, spacing 0.8 mm. 2) Intera 1.5T (Philips Healthcare, Netherlands), TR/TE 25/6.90 ms, FOV 180 × 180 mm, slice thickness 1.3 mm, spacing 0.65 mm. 3) Vantage Elan 1.5T (Canon Medical Systems, Japan), TR/TE 28/6.8 ms, FOV 200 × 200 mm, slice thickness 1.0 mm, spacing 0.5 mm. Maximum intensity projection (MIP) images were generated to optimize stenosis detection. ICAS was assessed in the major intracranial arterial segments, including the intracranial internal carotid artery (ICA), the M1 segment of the middle cerebral arteries, the A1 or A2 segment of the anterior cerebral arteries, the basilar artery, the V4 segment of the vertebral arteries, and the P1 or P2 segment of the posterior cerebral arteries. Stenosis was evaluated on both sides for paired vessels and defined as ≥50% luminal narrowing on time-of-flight (TOF) MRA, consistent with previous epidemiological and clinical studies.^18^ MRA images were independently reviewed by two board-certified neurologists who were blinded to clinical and laboratory data. When discrepancies occurred, a third neurologist (a board-certified consultant vascular neurologist) adjudicated the findings, and the final diagnosis was determined by consensus. Asymptomatic ICAS was defined as the presence of intracranial arterial stenosis without any history of stroke or TIA.

### 2.3. Dietary assessment

Nutritional status was assessed using a self-administered food frequency questionnaire (FFQ) for application in community-dwelling older Japanese adults. The FFQ is a booklet-style questionnaire that includes illustrations and photographs depicting portion sizes to facilitate completion by older adults. Assistance from family members was allowed, and all responses were checked by trained staff independent of the research team for completeness and internal consistency. The FFQ evaluates the habitual consumption of 233 food items and beverages over the preceding month and enables estimation of intake for 23 food groups and 160 nutrients. In this analysis, we examined total energy intake; major macronutrients (fat, protein, and carbohydrate); selected minerals (sodium and potassium); and dietary fiber. Energy intake was analyzed as a crude value, while all other nutrient intakes were energy-adjusted using the residual method as proposed by Willett and Stampfer.^19^ The validity and reproducibility of this FFQ have been examined in an ongoing validation study within the JPSC-AD cohort, and a summary report is publicly available from Kyushu University.^20^

### 2.4. Statistical analysis

Continuous variables are presented as mean ± standard deviation or median (interquartile range), as appropriate, based on their distribution, and categorical variables as number (percentage). Univariate comparisons of clinical characteristics were performed using the chi-square or Fisher’s exact test for categorical variables and the Student’s t-test or Mann–Whitney U test for continuous variables. For nutritional exposures, crude nutrient intakes (g/day or mg/day) were compared using the Mann–Whitney U test. Nutrient intakes were then energy-adjusted using the residual method, in which each nutrient was regressed on total energy intake and the unstandardized residuals were used for all regression analyses. Energy-adjusted nutrient variables were categorized into quartiles to account for non-normal distributions and facilitate interpretation. Odds ratios (ORs) and 95% confidence intervals (CIs) for ICAS across quartiles were estimated using logistic regression, with the lowest quartile serving as the reference. Three multivariable models were constructed: Model 1 adjusted for age and sex; Model 2 additionally adjusted for hypertension, dyslipidemia, and diabetes mellitus; and Model 3 further adjusted for body mass index (BMI), smoking status, and alcohol consumption. A two-tailed P < 0.05 was considered statistically significant. If multiple nutrients originate from overlapping food sources were significantly associated with ICAS, additional analyses were conducted to assess potential mutual confounding and collinearity between these nutrients. First, both energy-adjusted residual variables (continuous) for these nutrients were simultaneously entered into the multivariable logistic regression models to evaluate whether the association of either nutrient with ICAS was affected by adjustment for the other. Second, collinearity was evaluated using linear regression models including these nutrient residual variables, from which variance inflation factors (VIF) and tolerance values were derived. A VIF >5 was considered indicative of substantial multicollinearity. Analyses were performed using IBM SPSS Statistics, version 27.0.1 (IBM Corp., Armonk, NY, USA).

## 3. Results

### 3.1. Participant characteristics

Of the 962 community-dwelling older adults enrolled in the YAHABA study, 43 participants could not be evaluated by brain MRI. After excluding two participants with inadequate vascular imaging on MRA, 57 with a history of ischemic stroke, and 10 with missing dietary or clinical data, a total of 850 participants were included in the final analysis (Figure 1).

**Figure 1.**
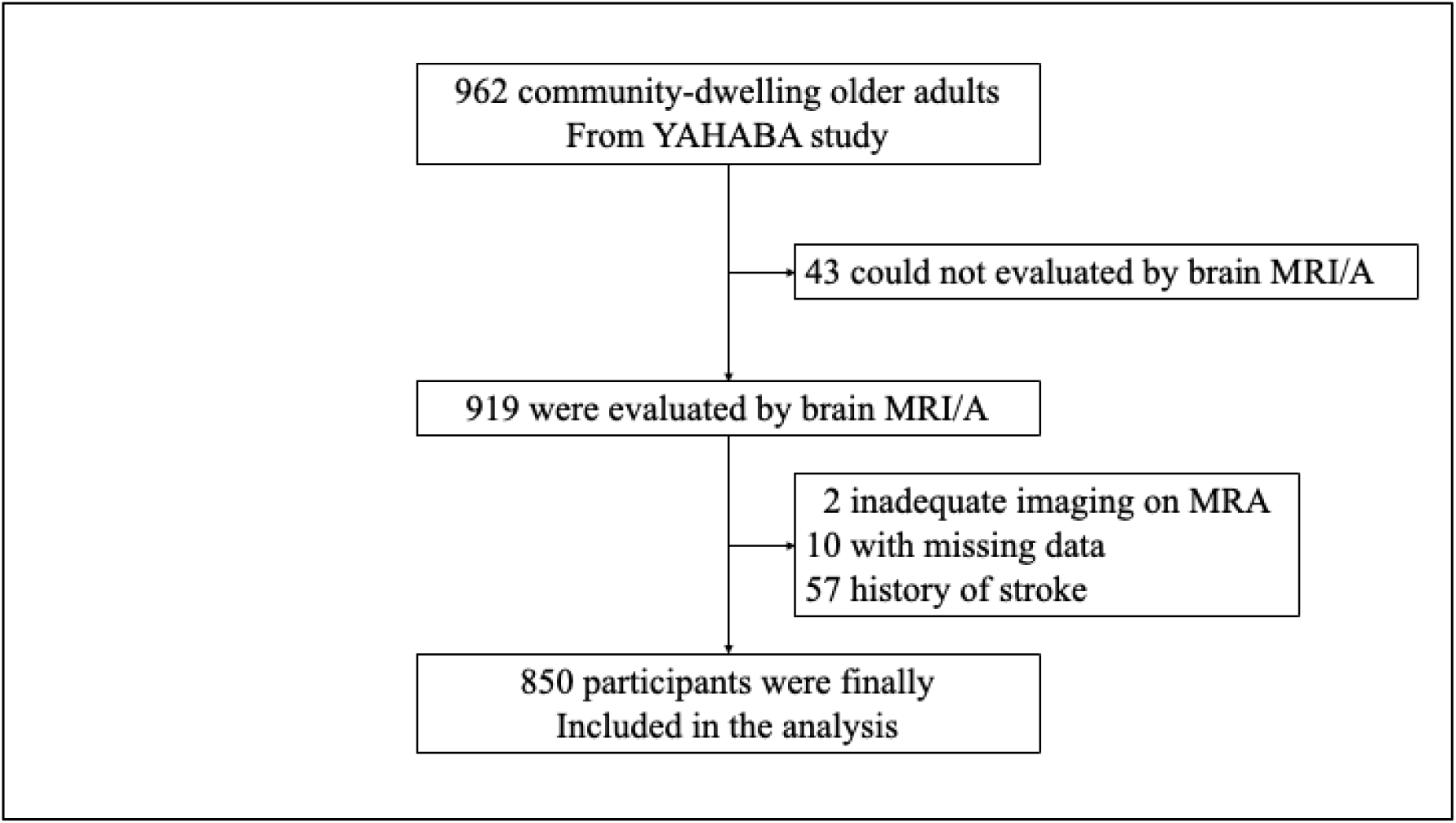
Flow chart of participant selection. A total of 850 participants were included in the final analysis. YAHABA study, Yahaba Active Aging and Healthy Brain study; MRI/A, magnetic resonance imaging/angiography.

Among them, ICAS was identified in 135 individuals (15.9%). Baseline characteristics and univariate comparisons between participants with and without ICAS are shown in Table 1. Participants with ICAS were significantly older than those without (75.8 ± 7.1 vs. 72.9 ± 6.3 years, P = 0.01). The prevalence of hypertension (69.6% vs. 51.7%, P < 0.01) and diabetes mellitus (22.2% vs. 14.3%, P = 0.02) was significantly higher in participants with ICAS than in those without. Systolic and diastolic blood pressure levels did not significantly differ between these groups. Renal function, assessed by eGFR, was significantly lower in participants with ICAS compared with those without (66.7 ± 12.3 vs. 69.8 ± 10.5 mL/min/1.73 m², P = 0.01). LDL cholesterol levels were comparable between the groups (111.7 ± 29.5 vs. 112.2 ± 28.4 mg/dL, P = 0.80), whereas HDL cholesterol was numerically lower in the ICAS group, although this difference was not statistically significant (58.2 ± 14.0 vs. 60.7 ± 15.6 mg/dL, P = 0.09). HbA1c levels were also comparable (5.8 ± 0.7% vs. 5.7 ± 0.7%, P = 0.28). There were no significant differences between the groups with respect to sex, body mass index, smoking status, alcohol consumption, or dyslipidemia.

**Table 1.** Subject demographics.

|  |  | Total | Without ICAS | With ICAS | p value |
| --- | --- | --- | --- | --- | --- |
| n |  | 850 | 715 | 135 |  |
| Age (year) |  | 73.4 ± 6.5 | 72.9 ± 6.3 | 75.8 ± 7.1 | 0.01 |
| Male, n (%) |  | 363 (42.4) | 308 (42.7) | 55 (40.7) | 0.67 |
| Smoking, n (%) | Current | 176 (20.6) | 150 (20.8) | 26 (19.3) | 0.34 |
|  | Former | 91 (10.6) | 81 (11.2) | 10 (7.4) |  |
|  | Never | 589 (68.8) | 490 (68.0) | 99 (73.3) |  |
| Drinking, n (%) |  | 75 (8.8) | 59 (8.2) | 16 (11.9) | 0.25 |
| BMI (kg/m<sup>2</sup>) |  | 24.2 ± 3.5 | 24.1 ± 3.6 | 24.5 ± 3.4 | 0.44 |
| Hypertension, n (%) |  | 465 (54.3) | 371 (51.7) | 94 (69.6) | <0.001 |
| Dyslipidemia, n (%) |  | 548 (64.0) | 463 (64.4) | 85 (63.0) | 0.75 |
| Diabetes Mellitus, n (%) |  | 133 (15.5) | 103 (14.3) | 30 (22.2) | 0.02 |
| SBP (mmHg) |  | 141.0 ± 17.0 | 140.5 ± 16.8 | 143.4 ± 17.7 | 0.37 |
| DBP (mmHg) |  | 79.0 ± 10.3 | 79.0 ± 10.3 | 78.8 ± 10.2 | 0.79 |
| eGFR (mL/min/1.73 m<sup>2</sup>) |  | 69.4 ± 10.9 | 69.8 ± 10.5 | 66.7 ± 12.3 | 0.01 |
| LDL-C (mg/dL) |  | 112.1 ± 28.6 | 112.2 ± 28.4 | 111.7 ± 29.5 | 0.80 |
| HDL-C (mg/dL) |  | 60.3 ± 15.4 | 60.7 ± 15.6 | 58.2 ± 14.0 | 0.09 |
| HbA1c (%) |  | 5.7 ± 0.7 | 5.7 ± 0.7 | 5.8 ± 0.7 | 0.28 |

Data are presented as mean ± standard deviation or number (percentage).
Comparisons between groups were performed using the Student's t-test for continuous variables and the chi-square test for categorical variables.
ICAS, intracranial atherosclerotic stenosis; n, number; BMI, body mass index; SBP, systolic blood pressure; DBP, diastolic blood pressure; eGFR, estimated glomerular filtration rate; LDL-C, low-density lipoprotein cholesterol; HDL-C, high-density lipoprotein cholesterol; HbA1c, glycated hemoglobin..

### 3.2. Nutritional status and ICAS

Unadjusted univariate comparisons of nutrient intakes between participants with ICAS and those without are presented in Table 2. Dietary fiber intake was significantly lower in participants with ICAS compared to those without (14.5 [11.4-17.4] vs. 15.5 [12.2-18.8] g/day, P = 0.03). Potassium intake was numerically lower in participants with ICAS than in those without (2992.9 [2346.6–3924.9] vs. 3053.8 [2524.6–3668.0] mg/day, P = 0.06). In contrast, there was no significant difference in total energy intake, macronutrients (fat, protein, and carbohydrate), and sodium intake between these groups.

**Table 2.** Comparison of nutrient intakes between participants with and without asymptomatic ICAS.

| Nutrients | Without ICAS | With ICAS | p value |
| --- | --- | --- | --- |
| n | 715 | 135 |  |
| Total energy (kcal/day) | 1805.5<br>(1585.0-2068.8) | 1759.8<br>(1532.2-2278.9) | 0.31 |
| Fat (g/day) | 53.6<br>(43.3-65.7) | 52.6<br>(43.8-74.9) | 0.58 |
| Protein (g/day) | 70.8<br>(59.8-84.2) | 69.7<br>(59.1-84.3) | 0.55 |
| Carbohydrates (g/day) | 237.8<br>(208.9-270.2) | 233.5<br>(201.8-263.3) | 0.55 |
| Dietary fiber (g/day) | 15.5<br>(12.2-18.8) | 14.5<br>(11.4-17.4) | 0.03 |
| Sodium (mg/day) | 4210.0<br>(3439.5-5192.1) | 4202.4<br>(3273.9-6109.3) | 0.58 |
| Potassium (mg/day) | 3053.8<br>(2524.6-3668.0) | 2992.9<br>(2346.6-3924.9) | 0.06 |

Values are presented as median (interquartile range). P values were derived from the Mann–Whitney U test. Nutrient intakes are unadjusted. ICAS, intracranial atherosclerotic stenosis; n, number.

Univariate models employing energy-adjusted nutrient intakes using the residual method are presented in Table 3. Participants in the highest quartile of dietary fiber intake had significantly lower odds of ICAS compared to those in the lowest quartile (OR, 0.54; 95% CI, 0.32–0.93; P for trend = 0.03). Participants in the highest quartile of potassium intake also had lower odds of ICAS compared to those in the lowest quartile (OR, 0.56; 95% CI, 0.32–0.97), although the trend was borderline significant (P for trend = 0.05). There were no significant associations for other nutrients, including fat, protein, carbohydrate, and sodium.

**Table 3.**
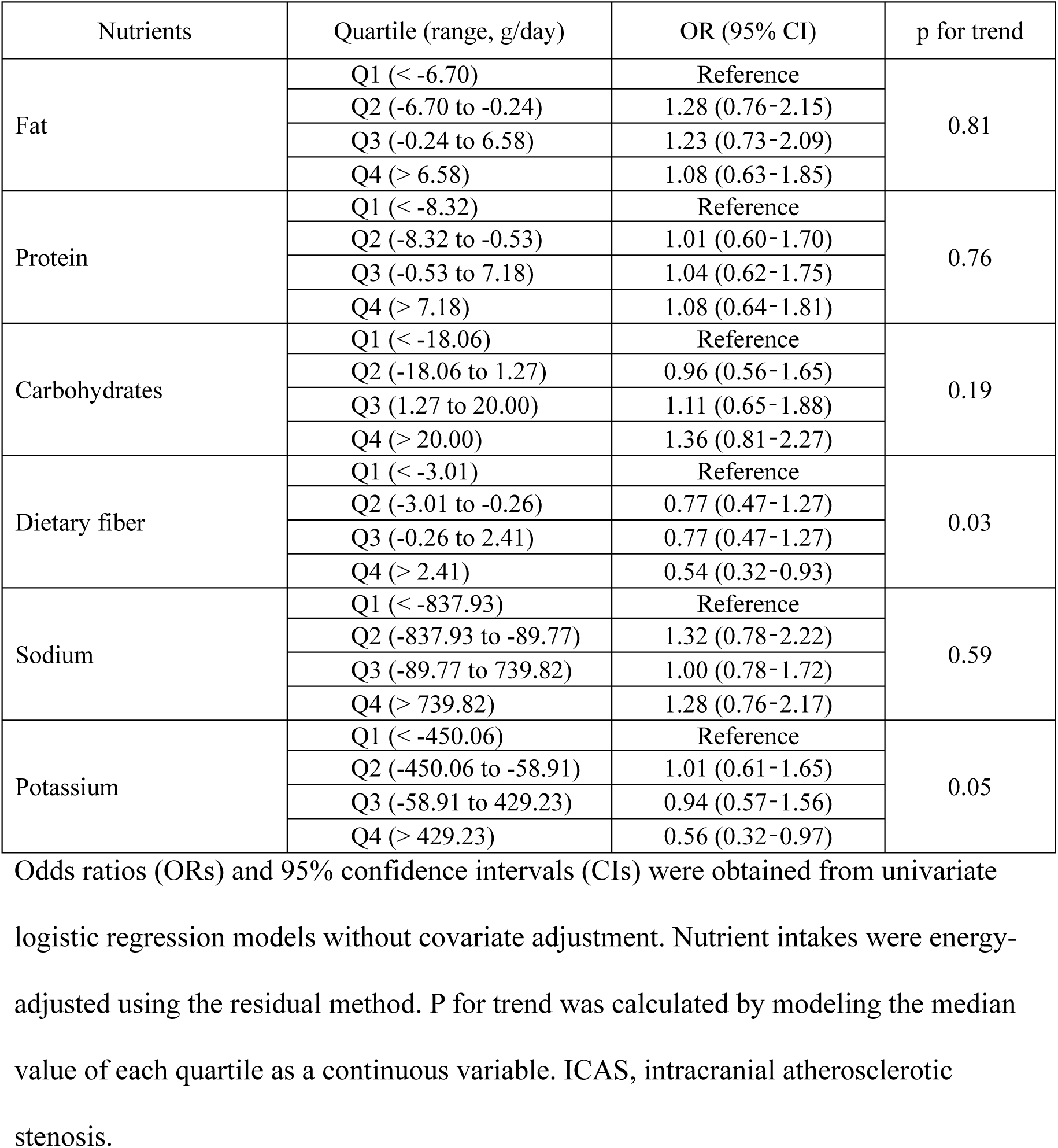
Univariate associations between energy-adjusted nutrient intakes and asymptomatic ICAS.

### 3.3. Multivariable analysis between energy-adjusted nutrient intakes and ICAS

Multivariable logistic regression analysis between energy-adjusted nutrient intakes and asymptomatic ICAS across quartiles are presented in Table 4 and Figure 2. Dietary fiber intake was consistently associated with lower odds of ICAS across all adjustment models. In the fully adjusted model (Model 3), participants in the highest quartile of dietary fiber intake had a significantly lower odds of ICAS than those in the lowest quartile (OR 0.45, 95% CI 0.26–0.80; P for trend = 0.01). Potassium intake also showed a similar association with ICAS. In Model 3, the highest quartile of potassium intake was associated with a significantly lower odds of asymptomatic ICAS compared with the lowest quartile (OR 0.49, 95% CI 0.27–0.89; P for trend = 0.03) (Figure 2). In contrast, there were no significant associations with fat, protein, carbohydrate, or sodium intake in any multivariable model (all P for trend > 0.25).

**Figure 2.**
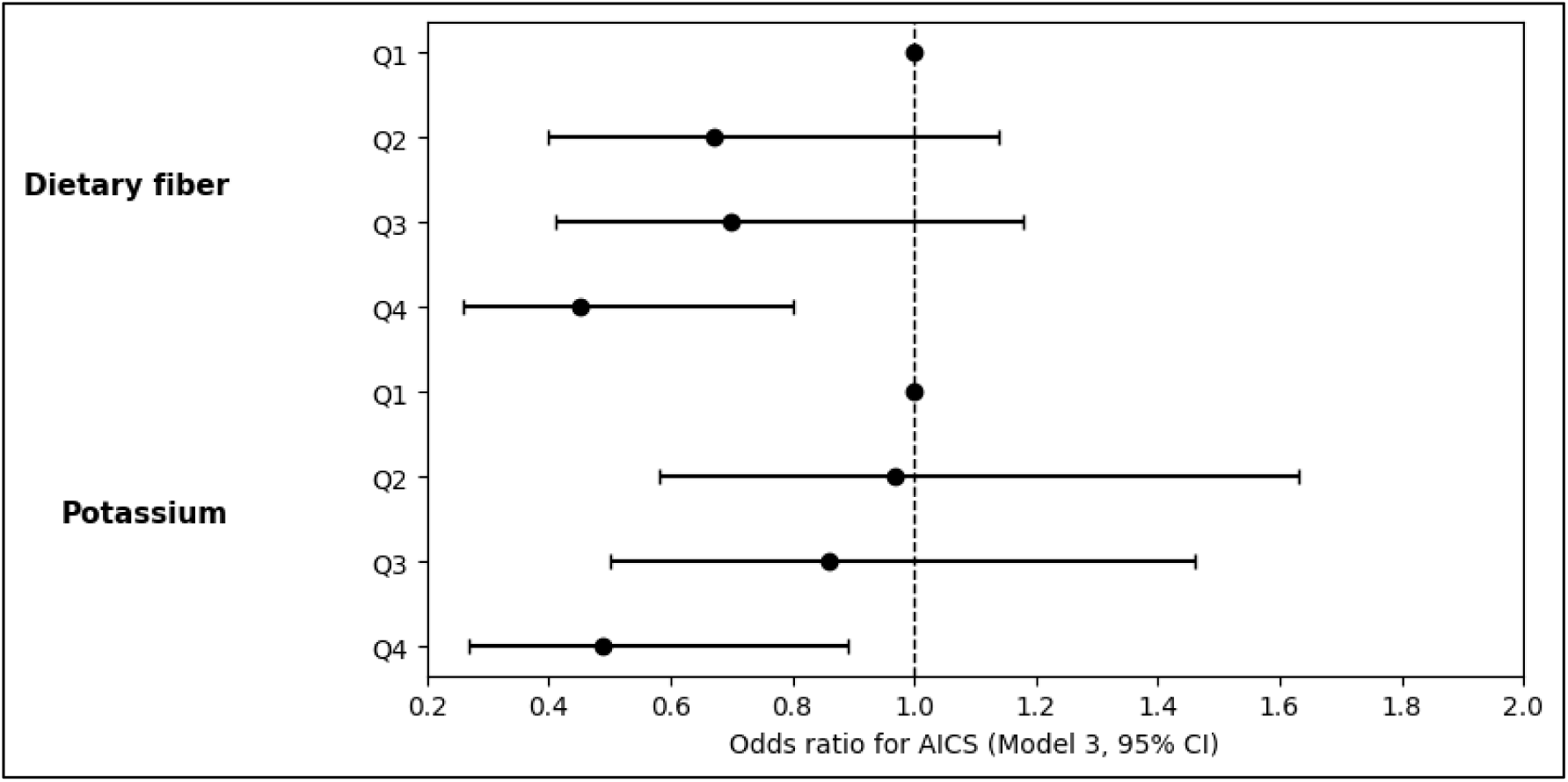
Forest plot of the association between dietary fiber and potassium intakes and asymptomatic ICAS. Multivariable-adjusted odds ratios (ORs) and 95% confidence intervals (CIs) for asymptomatic intracranial atherosclerotic stenosis (ICAS) according to quartiles of dietary fiber and potassium intakes. Nutrient intakes were energy-adjusted using the residual method and categorized into quartiles (Q1–Q4), with the lowest quartile (Q1) serving as the reference. ORs were estimated using logistic regression adjusted for age, sex, hypertension, dyslipidemia, diabetes mellitus, body mass index, smoking status, and alcohol consumption. The dashed vertical line indicates an OR of 1.0.

**Table 4.**
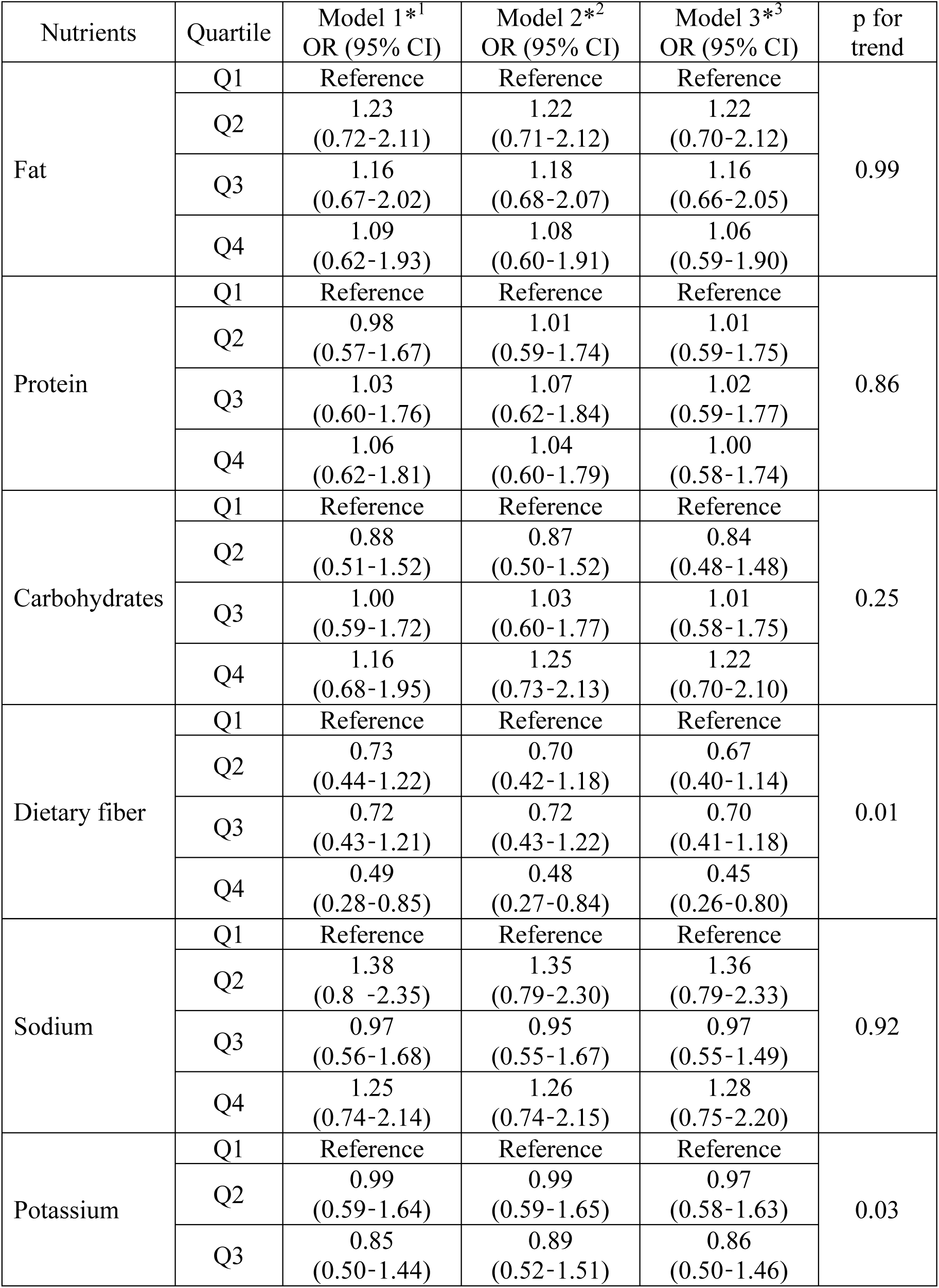
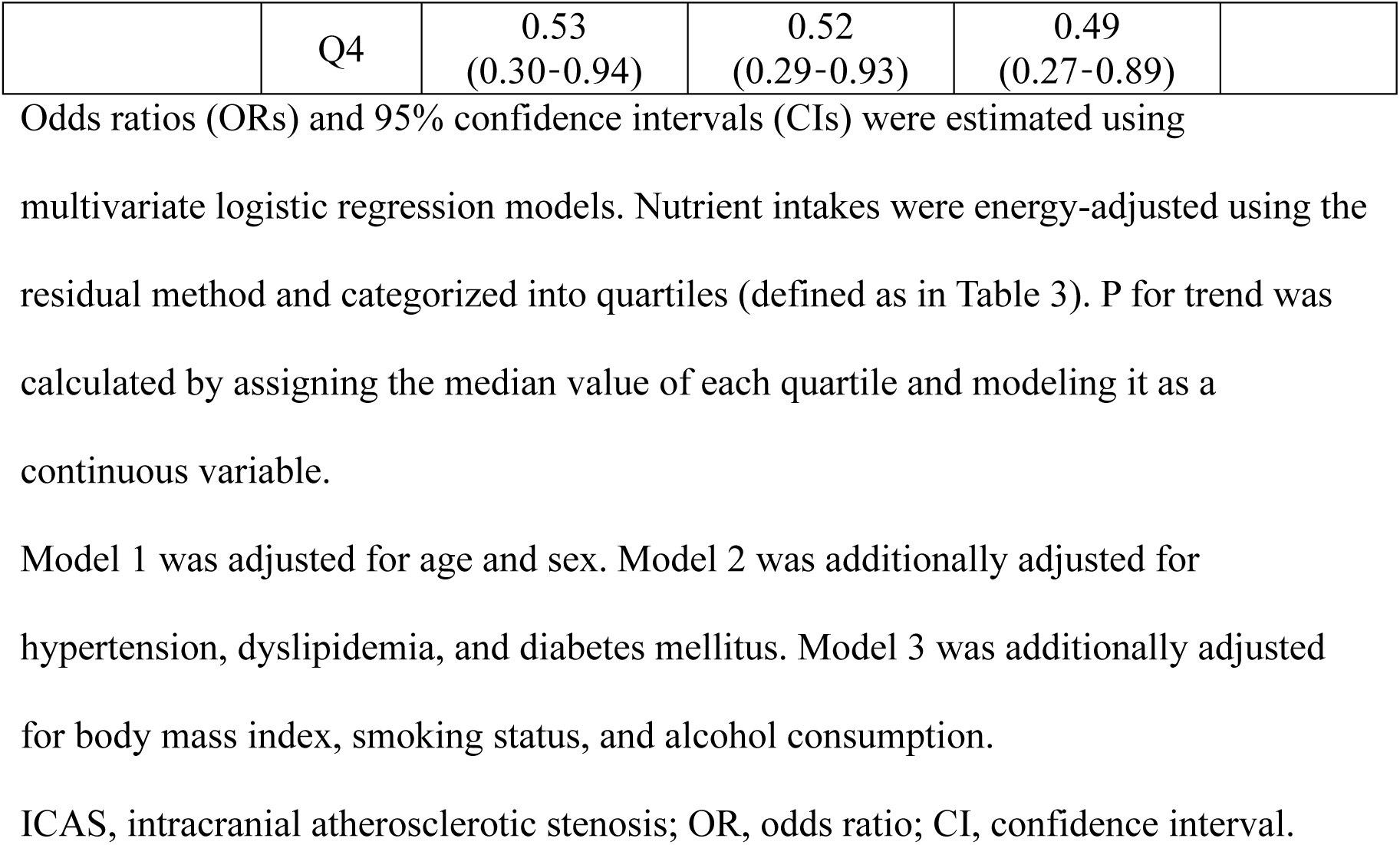
Multivariate-adjusted associations between energy-adjusted nutrient intakes and asymptomatic ICAS.

### 3.4. Sensitivity analyses and assessment of collinearity

To explore whether dietary fiber and potassium exerted independent effects on ICAS, both nutrients were simultaneously entered into the fully adjusted logistic regression models using continuous energy-adjusted residual variables (scaled per 5 g/day for dietary fiber and per 100 mg/day for potassium). Under this specification, neither dietary fiber nor potassium remained statistically significant (dietary fiber: OR, 0.97 per 5 g increase; 95% CI, 0.62–1.51; potassium: OR, 0.96 per 100 mg increase; 95% CI, 0.91–1.02). Collinearity diagnostics revealed moderate correlation between dietary fiber and potassium intake, with a VIF of 4.16 for both variables.

## 4. Discussion

In this well-characterized community-based cohort, comprehensive dietary assessment with a validated food frequency questionnaire showed that higher dietary fiber intake was associated with lower odds of asymptomatic ICAS. Potassium intake demonstrated a similar inverse association, although the association was somewhat weaker and less consistent after accounting for collinearity with dietary fiber. These associations were consistent across multivariable models adjusted for established vascular risk factors. When dietary fiber and potassium were simultaneously included in the multivariable regression model as continuous variables, neither remained statistically significant, suggesting that the observed associations may be attributable to shared dietary sources.

Previous epidemiological studies have consistently reported inverse associations between dietary fiber intake and cardiovascular disease, ischemic stroke, and extracranial atherosclerosis.^14, 21, 22^ However, data specifically addressing intracranial arterial disease, particularly at the asymptomatic stage in community-dwelling populations, are limited. Several cardiometabolic pathways may underlie this association. Dietary fiber has been shown to modestly reduce blood pressure^23^ and LDL cholesterol^24^ and to improve glycemic control, including reductions in fasting blood glucose and HbA1c in individuals with type 2 diabetes.^25^ In addition, fermentation of dietary fiber produces short-chain fatty acids^26^ with anti-inflammatory effects on endothelial cells,^27^ and higher fiber intake attenuates postprandial glycemic excursions.^28^ Collectively, these mechanisms may contribute to endothelial protection beyond conventional risk factor modification.

Potassium intake also showed an association with asymptomatic ICAS in community-dwelling older adults, consistent with its established role in vascular health. Potassium intake has been shown to lower blood pressure^29^ and may also confer additional benefits on endothelial function.^30^ Nevertheless, direct evidence linking potassium intake to intracranial atherosclerosis remains sparse. In the present study, the association between potassium intake and asymptomatic ICAS was directionally consistent with that of dietary fiber but slightly weaker, suggesting that potassium intake may be associated with asymptomatic ICAS as part of a broader dietary pattern rather than as an isolated exposure.

Sensitivity analyses and assessment of collinearity suggested moderate collinearity between dietary fiber and potassium. Dietary fiber and potassium often share dietary sources, as foods rich in dietary fiber—such as vegetables, fruits, seaweed, and mushrooms—also contribute substantially to potassium intake. Thus, the attenuation observed in the simultaneous model should not be interpreted as evidence against the relevance of either nutrient. Rather, it highlights the inherent difficulty of disentangling individual nutrient effects when they are embedded within common plant-based dietary patterns^25^. In our findings, the primary quartile analyses, which are less sensitive to continuous-scale collinearity, demonstrated a clear inverse association between higher fiber intake and asymptomatic ICAS. Taken together with the observed collinearity between dietary fiber and potassium, these findings suggest that the associations may reflect underlying plant-based dietary patterns rather than the isolated effects of a single nutrient.

Several limitations should be acknowledged. The cross-sectional design precludes causal inference, and reverse causation cannot be excluded. Dietary intake was assessed using a FFQ and therefore could be subject to measurement error. Although we examined collinearity between dietary fiber and potassium, residual confounding by other correlated dietary components or unmeasured lifestyle factors remains possible. In addition, this study was conducted in a single geographic region and included only older Japanese adults, which may limit the generalizability of the findings to other populations. Finally, TOF-MRA allowed assessment of luminal narrowing but could not provide information on plaque composition. Future longitudinal or interventional studies are warranted to determine whether increasing dietary fiber and potassium intake can modify the progression of intracranial atherosclerosis or reduce the risk of cerebrovascular events.

## Conclusion

Higher dietary fiber intake was inversely associated with asymptomatic ICAS in community-dwelling older Japanese adults. Potassium intake also showed an inverse association, although this relationship was less consistent. These findings suggest that dietary patterns characterized by higher consumption of plant-derived foods may be associated with a lower prevalence of asymptomatic ICAS.

## Data Availability

The data that support the findings of this study are available from the corresponding author upon reasonable request, subject to approval by the institutional ethics committee.

## Acknowledgments

The authors wish to thank all older adults who participated in the study, as well as the staff of Yahaba Town Hall, for their cooperation. The authors would like to express their sincere gratitude to Researchers Yasuo Terayama MD, PhD, Hisashi Yonezawa MD, PhD, and Junko Takahashi MD, PhD for their valuable contributions to the establishment of this study. The authors also thank Ms. Akie Sakamoto and Ms. Kuniko Watanabe for their dedicated assistance with the clinical examinations.

## Non-standard Abbreviations and Acronyms

ICAS: intracranial atherosclerotic stenosis
TIA: transient ischemic attack
MRA: magnetic resonance angiography
YAHABA: Yahaba Active Aging and Healthy Brain
JPSC-AD: Japan Prospective Studies Collaboration for Aging and Dementia
MRI: magnetic resonance imaging
MIP: maximum intensity projection
ICA: internal carotid artery
MCA: middle cerebral artery
ACA: anterior cerebral artery
PCA: posterior cerebral artery
BA: basilar artery
V4: V4 segment of vertebral artery
P1: P1 segment of posterior cerebral artery
P2: P2 segment of posterior cerebral artery
FFQ: food frequency questionnaire
eGFR: estimated glomerular filtration rate
HbA1c: hemoglobin A1c
HDL: high-density lipoprotein
LDL: low-density lipoprotein
BMI: body mass index
OR: odds ratio
CI: confidence interval
VIF: variance inflation factor
TOF: time-of-flight
TR: repetition time
TE: echo time
FOV: field of view

